# CD14 Positive Extracellular Vesicles in Broncho-Alveolar Lavage Fluid as a New Biomarker of Acute Respiratory Distress Syndrome

**DOI:** 10.1101/2021.09.25.21264053

**Authors:** Rahul Y. Mahida, Joshua Price, Sebastian T. Lugg, Hui Li, Dhruv Parekh, Aaron Scott, Paul Harrison, Michael A. Matthay, David R. Thickett

## Abstract

Recent studies have indicated that extracellular vesicles (EV) may play a role in the pathogenesis of Acute Respiratory Distress Syndrome (ARDS). EV have been identified as potential biomarkers of disease severity and prognosis in other pulmonary diseases. We sought to characterize the EV phenotype within ARDS patient broncho-alveolar lavage fluid (BAL), and to determine whether BAL EV could be utilized as a potential biomarker in ARDS. BAL was collected from sepsis patients with and without ARDS, and from esophagectomy patients post-operatively (of whom a subset later developed ARDS during hospital admission). BAL EV were characterized with regards to size, number and cell of origin. Sepsis patients with ARDS had significantly higher numbers of CD14^+^/CD81^+^ monocyte-derived BAL EV than sepsis patients without ARDS (p=0.015). However, the converse was observed in esophagectomy patients who later developed ARDS (p=0.003). Esophagectomy patients who developed ARDS also had elevated CD31^+^/CD63^+^ and CD31^+^/CD81^+^ endothelial-derived BAL EV (p≤0.02) compared to esophagectomy patients who did not develop ARDS. Further studies are required to determine whether CD31^+^ BAL EV may be a predictive biomarker for ARDS in esophagectomy patients. CD14^+^/CD81^+^ BAL EV numbers were significantly higher in those patients with sepsis-related ARDS who died during the 30 days following ICU admission (p=0.027). Thus, CD14^+^/CD81^+^ BAL EV are a potential biomarker for disease severity and mortality in sepsis-related ARDS. These findings provide the impetus to further elucidate the contribution of these EV to ARDS pathogenesis.

## INTRODUCTION

Acute Respiratory Distress Syndrome (ARDS) is a hyper-inflammatory pulmonary disorder, which most commonly develops following sepsis. Neutrophilic inflammation and alveolar-capillary barrier damage lead to alveolar edema and refractory hypoxia, requiring prolonged mechanical ventilation (1). Mortality remains high at 35-45% (2). Survivors have long-term morbidity and increased risk of developing lung fibrosis. There is an urgent need to identify novel therapeutic targets for ARDS, especially due to the evolving SARS-CoV-2 pandemic in which ARDS is the main cause of mortality (3).

Extracellular vesicles (EV) are membrane-bound anuclear structures which constitute an inter-cellular communication mechanism (4). EV allow targeted transfer of diverse biologic cargo (including mitochondria, RNA, cytokines) between different cell types (5). Uptake of EV can alter gene expression within host cells (6). The tetraspanins CD9, CD63 and CD81 are transmembrane proteins commonly expressed on EV, and can be used to isolate EV in acellular biofluids (7). There are two main subtypes of EV: exosomes and microvesicles. Exosomes are 30-150nm in diameter and form by fusion of multi-vesicular bodies with the plasma membrane. Microvesicles are 100-1000nm in diameter and form by outward blebbing of the plasma membrane (4). With overlap in size and common markers (e.g. tetraspanins), differentiating exosomes and microvesicles is challenging, however Endosomal Sorting Complex Required for Transport (ESCRT) proteins such as syntenin are candidate internal cargo markers specific to exosomes (8).

Recent evidence indicates that EV play an important role in ARDS pathogenesis by promoting persistent inflammation. Pathogenic EV are released when human *ex-vivo* perfused lungs are injured with *E. coli*; isolation of these EV and administration to uninjured human lungs can also induce inflammatory lung injury (9). Murine models of lipopolysaccharide lung injury have shown that EV transfer of microRNA cargo (e.g. miR-466) to alveolar macrophages can increase inflammatory cytokine release (10). Recent studies have indicated that CD14^+^ BAL EV may be a potential biomarker for disease activity in COPD (11).

We previously showed that alveolar macrophages isolated from patients with sepsis-related ARDS have impaired efferocytosis (ability to clear apoptotic cells) compared to control ventilated sepsis patients without ARDS (12). Alveolar macrophage efferocytosis negatively correlated with alveolar neutrophil apoptosis, and broncho-alveolar lavage (BAL) cytokines IL-8 and IL-1α. Impaired efferocytosis was associated with increased 30-day mortality and duration of mechanical ventilation, indicating that this alveolar macrophage functional defect contributes to ARDS pathogenesis (12). Treatment of healthy alveolar macrophages with BAL from ARDS patients also impairs efferocytosis and downregulates Rac1 expression (which causes cytoskeletal rearrangement allowing apoptotic cell engulfment), reproducing the same functional defect observed in ARDS (13). However, the causative immunomodulatory factors within ARDS patient BAL remain to be identified. Identification of differentially expressed EV populations within ARDS BAL may guide further investigation into the mechanism of this alveolar macrophage functional defect

In this study, the objective was characterize the EV phenotype in the BAL of patients with sepsis-related ARDS and to compare this EV population to that in control sepsis patients and in post-esophagectomy patients (a subset of whom later developed ARDS). We also aimed to determine whether BAL EV could be utilized as a potential biomarker in sepsis-related ARDS.

## MATERIALS AND METHODS

### Ethical Approval

As previously described (12), ethical approval was obtained to recruit ventilated sepsis patients with and without ARDS (REC 16/WA/0169). Ethical approval was also obtained to recruit patients undergoing transthoracic esophagectomy for esophageal carcinoma (REC 12/WM/0092) in the VINDALOO trial (14, 15). For patients who lacked capacity, permission to enroll was sought from a personal legal representative in accordance with the UK Mental Capacity Act (2005). For patients with capacity, written informed consent was obtained from the patient.

### Patient recruitment and broncho-alveolar lavage

Invasively ventilated adult sepsis patients with and without ARDS were recruited from the intensive care unit of the Queen Elizabeth Hospital Birmingham, U.K. from December 2016 – February 2019 and BAL was collected as previously described, within 48 hours of initiation of mechanical ventilation (12). Patient demographic and physiological details are in this prior publication. In the VINDALOO trial (15), patients underwent post-operative bronchoscopy and BAL following esophagectomy. Following unblinding of the study, those patients who received placebo were identified; BAL samples from the 4 patients who developed ARDS during their post-operative course and 8 patients who did not develop ARDS were analyzed in this study. For all patients, sterile, isotonic saline (2 × 50ml aliquots) was instilled into a sub-segmental bronchus of the lingula or right middle lobe. Recovery ranged between 20 – 60 ml and did not differ between patient groups. BAL fluid collection was standardized across all patients in keeping with European guidelines (16).BAL fluid was rendered acellular by centrifuging at 500g for 5 minutes then stored at -80°C in aliquots prior to use in this study. Differential cell count was performed on cells isolated from BAL.

### BAL Extracellular Vesicle characterization

A single-particle interferometric reflectance image sensing platform was used (Exoview R100 reader, NanoView Biosciences, USA) to detect and characterize EV size, number and surface marker expression within BAL samples (17). The Exoview platform allows quantification and phenotyping of EV ≥50nm in diameter (7). EV numbers in BAL were measured using ExoView Tetraspanin kits (NanoView Biosciences) according to manufacturer’s instructions. Each spot on an Exoview Tetraspanin chip is an independent test, interacting with only a small volume surrounding itself. Accounting for volume and spot size, a normalized particle number can be generated and when multiplied by the dilution factor and a correction factor of 10^4^, data can be expressed as EV/ml. Each spot on an Exoview Tetraspanin chip has the capacity to bind up to 5000 EVs before becoming saturated. According to the manufacturer’s instructions, at least a 1:1 dilution of any biofluid with incubation solution is required prior to incubation on Exoview Tetraspanin chips. Preliminary studies had found that use of dilutions lower than 1:20 resulted in chip saturation, with the number of EVs exceeding the chip binding capacity. BAL samples were therefore diluted 1:20 with incubation solution to prevent chip saturation. 35 µl of diluted BAL was loaded on the ExoView Tetraspanin chips and incubated for 16 hours in a sealed, humidified 24-well plate at room temperature to allow EV binding. Each chip contained spots printed with mouse anti-human antibodies against the tetraspanins CD63 (clone H5C6), CD81 (clone JS81) and CD9 (clone H19a) which enabled EV capture. Chips also contained spots with mouse IgG1*κ* matching isotype antibodies, used as a control for non-specific EV binding. Chips were washed thrice with incubation solution; after each wash plates were shaken at 500rpm (LSE digital microplate shaker, Corning, USA). Chips were then stained with a fluorescent antibody cocktail for 1 hour at room temperature in the dark to characterize EV surface markers. Fluorophore-conjugated mouse anti-human antibodies included: CD14-Allophycocyanin (1:100 dilution, clone 61D3, Invitrogen), CD206-Phycoerythrin (1:100 dilution, clone 19.2, Invitrogen), EpCam-AlexaFluor488 (1:400 dilution, clone 94C, BioLegend), CD66b-AlexaFluor647 (1:200 dilution, clone G10F5, BD Biosciences), CD31-Phycoerythrin (1:100 dilution, clone WM59, Invitrogen), and CD41-AlexaFluor488 (1:400 dilution, clone HIP8, BioLegend). Chips were washed thrice again, with a final wash in distilled water, before being imaged using the ExoView R100 reader and nScan2 v2.76 software. Data were analyzed using NanoViewer v2.82, utilizing mouse IgG capture spots as negative control (indicative of non-specific binding) for fluorescent gating, and EV sizing thresholds set at 50-200nm diameter. EV cell origin data were correlated with previously recorded clinical outcomes and immune cell functional parameters (12). Data from each tetraspanin capture spot (CD63, CD81 and CD9) were calculated by subtracting negative control values from IgG spots.

### Statistical Analysis

Data were analyzed using Prism 8 software (GraphPad, USA). Parametric data shown as mean and standard deviation. Non-parametric data shown as median and interquartile range. Differences between continuously distributed non-parametric data assessed using Mann-Whitney tests. Differences between three or more unpaired non-parametric data sets assessed using Kruskal-Wallis test by ranks. Two-tailed p-values of ≤0.05 were considered significant.

## RESULTS

The demographic and clinical characteristics of ARDS and control patients from the two cohorts analyzed are shown in **Table 1**. BAL samples were analyzed from 17 sepsis patients with ARDS, 14 sepsis patients without ARDS, 4 esophagectomy patients who later developed ARDS and 8 esophagectomy patients who did not develop ARDS.

**Table 1:** Patient demographic and clinical characteristics.

|  | Sepsis Patients |  | Esophagectomy Patients |  |
| --- | --- | --- | --- | --- |
|  | With ARDS<br>(n=17) | Without ARDS<br>(n=14) | Developed<br>ARDS<br>(n=4) | Did not<br>develop<br>ARDS (n=8) |
| **Demographics** |  |  |  |  |
| **Age in years:** | 59.2 (13.9) | 55.1 (16.3) | 63.3 (6.9) | 65.8 (7.9) |
| **Male Gender: n (%)** | 15 (71%) | 11 (65%) | 4 (100%) | 4 (50%) |
| **Ethnicity: Caucasian - n (%)** | 21 (100%) | 17 (100%) | 4 (100%) | 7 (88%) |
| **Smoking status: n (%)** |  |  |  |  |
| - Current | 7 (33%) | 5 (29%) | 1 (25%) | 0 (0%) |
| - Ex-smoker | 9 (43%) | 6 (35%) | 1 (25%) | 6 (75%) |
| - Never smoker | 2 (10%) | 1 (6%) | 2 (50%) | 2 (25%) |
| - Unknown | 3 (14%) | 5 (29%) | 0 (0%) | 0 (0%) |
| **ICU LoS in days:** | 23.0 | 12.0 | 9.5 | 4.5 |
| **median (IQR)** | (12.8 – 33.8) | (7.5 – 19.0) | (5 – 22.3) | (2.5 – 7.8) |
| **Hospital LoS in days:** | 33.0 | 24.0 | 22.0 | 12 |
| **median (IQR)** | (15.3 – 52.5) | (13.0 – 39.5) | (17.3 – 53.8) | (8.3 – 15.8) |
| **Inpatient Mortality: n (%)** | 7 (33.3%) | 3 (17.6%) | 1 (25%) | 0 (0%) |
| **BAL leukocyte count $\times 10^6$:** | 15.8 | 6.4 | 11.0 | 7.8 |
| **median (IQR)** | (7.4 – 31.3) | (3.8 – 27.0) | (5.9 – 18.6) | (2.1 – 10.5) |
| **% Neutrophils in BAL:** | 69.9 (21.5) | 49.0 (30.6) | 20 (20) | 9.5 (10.1) |

IQR = Interquartile range. LoS = Length of stay. ICU = Intensive care unit. Data are presented as mean (standard deviation) unless otherwise indicated.

BAL EV were characterized with regards to size, number and surface marker expression to indicate cellular origin. The size distribution of BAL EV did not differ across patient groups (**Figure 1A)**. Representative fluorescence images from the Exoview platform demonstrating EV surface marker expression and lack of non-specific binding are shown in **Figure 1B-E**.

**Figure 1:**
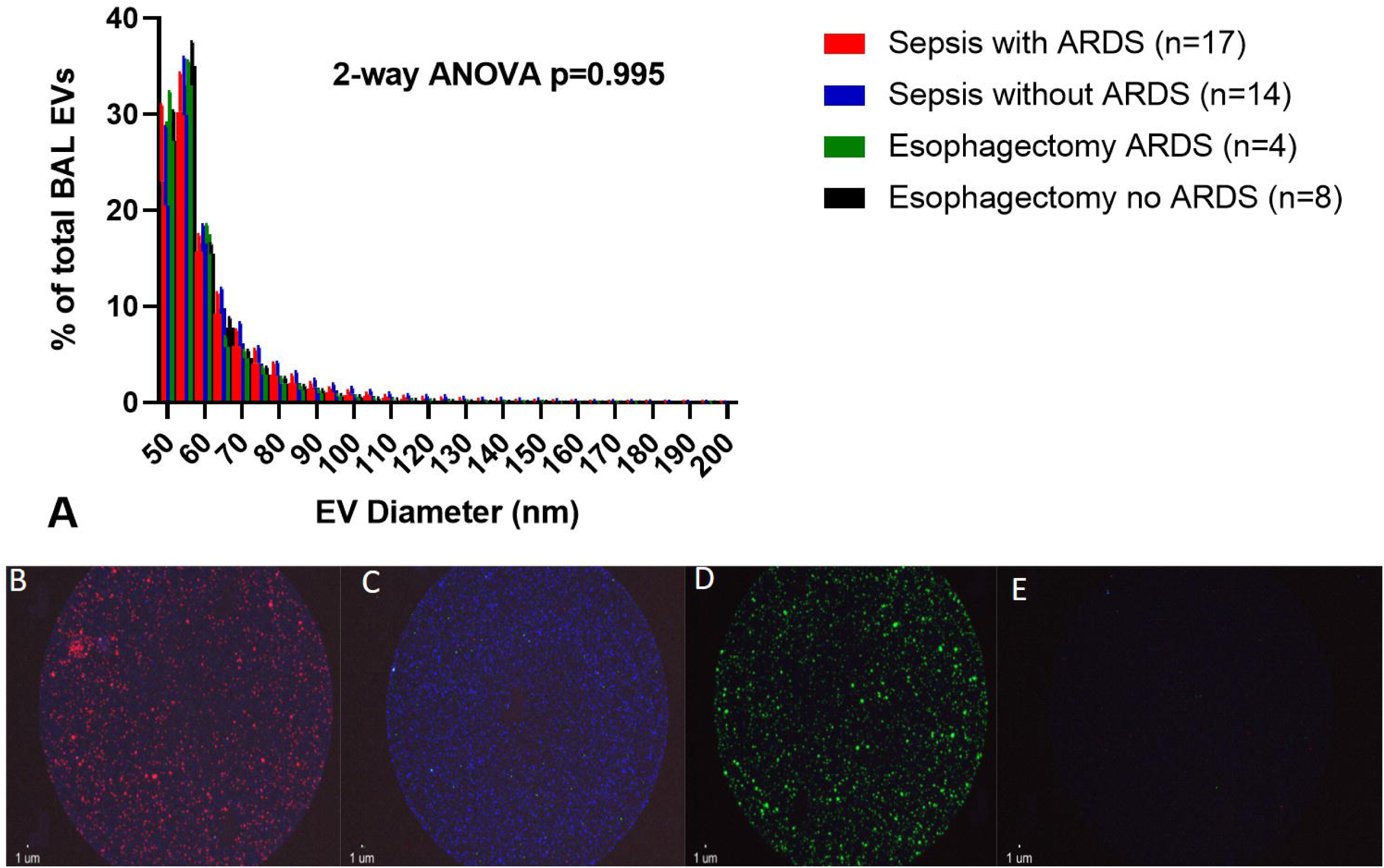
BAL EV size distribution and representative Exoview fluorescence images. **A**) BAL EV size distribution measured using single-particle interferometric reflectance image sensing (Exoview R100 platform). Statistical analysis by Kruskal-Wallis test, n≥4 in each group. There was no significant difference in EV size distribution between sepsis patients with ARDS, sepsis patients without ARDS, esophagectomy patients who developed ARDS and esophagectomy patients who did not develop ARDS (p=0.123). EV measured between 150-200nm are not shown in this graph as these accounted for <1% of total EV population in all three groups. Panels B-D show representative Exoview fluorescence images demonstrating EV surface marker expression. **B**: Detection of CD66b^+^ EV (AlexaFluor647) on CD63 tetraspanin spot. **C**: Detection of EpCam^+^ EV (AlexaFluor488) on CD9 tetraspanin spot. **D**: Detection of CD31^+^ EV (Phycoerythrin) on CD9 tetraspanin spot. **E**: Representative Mouse IgG spot negative control showing low non-specific EV binding in BAL samples.

Characterization studies showed that a greater number of monocyte-derived CD14^+^/CD81^+^ EV were present in the BAL of sepsis patients with ARDS compared to sepsis patients without ARDS (**Figure 2A**, medians 1.23 ×10^8^/ml vs 6.26 ×10^7^/ml, p=0.015). However in esophagectomy patients, those who subsequently developed ARDS had lower CD14^+^/CD81^+^ BAL EV counts compared to patients who did not develop ARDS (**Figure 2A**, medians 3.96 ×10^7^/ml vs 9.94 ×10^7^/ml, p=0.03). Endothelial-derived CD31^+^/CD63^+^ and CD31^+^/CD81^+^ BAL EV were elevated in esophagectomy patients who subsequently developed ARDS compared to those who did not develop ARDS (**Figure 2B**, p≤0.02). There were no differences in the numbers of CD66b^+^, CD41^+^, CD206^+^ and EpCam^+^ BAL EV between ARDS and control groups in both cohorts (**Figure 2C-F**).

**Figure 2:**
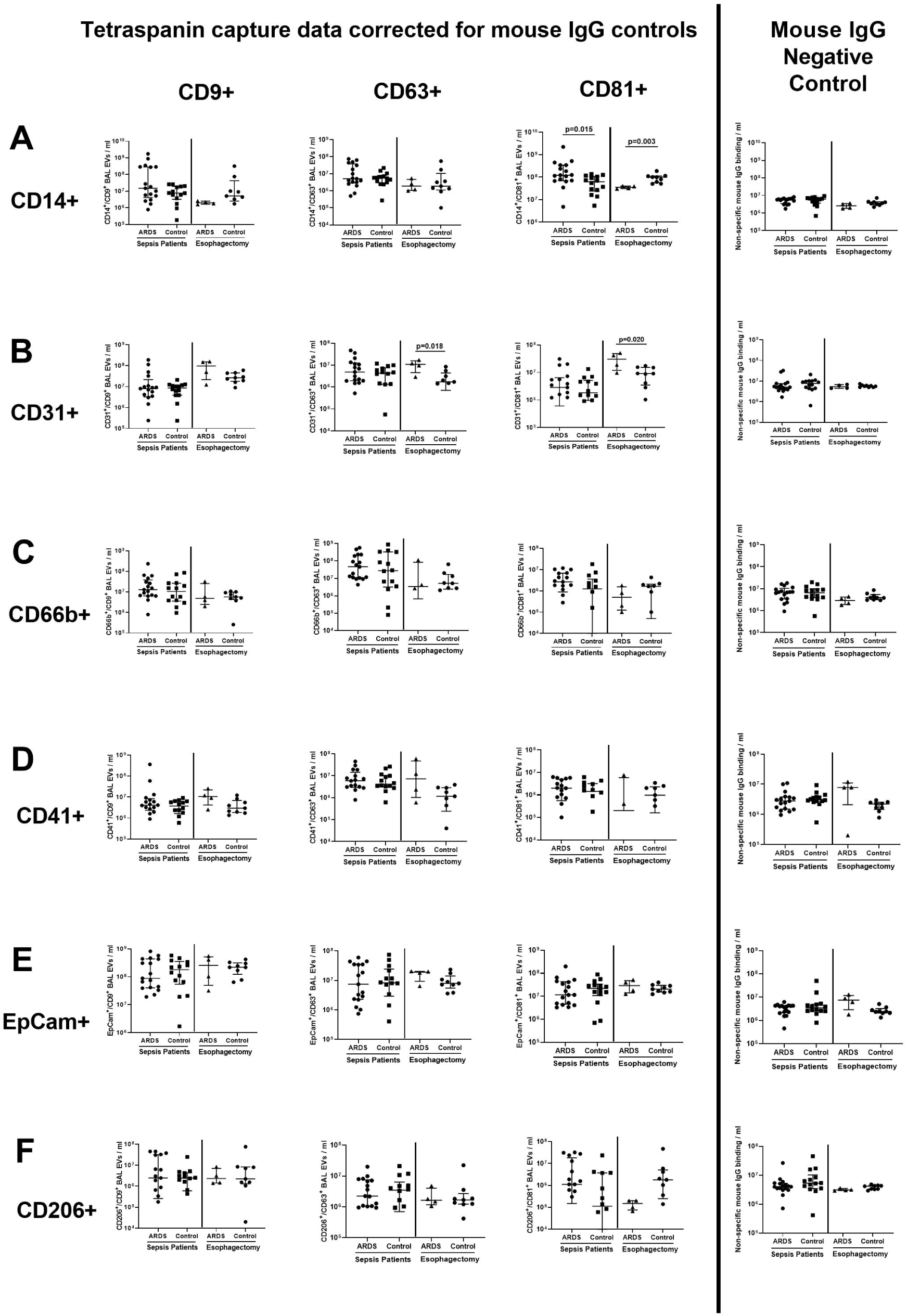
BAL EV surface marker expression in ARDS and control groups across sepsis and esophagectomy patient cohorts. BAL EV surface marker expression measured by tetraspanin antibody capture (CD9/63/81) and fluorescent antibody labelling via the Exoview R100 platform (EpCam for epithelial, CD66b for neutrophil, CD206 for alveolar macrophage, CD31 for endothelial, CD41 for platelet and CD14 for monocyte origin). All data expressed as median and inter-quartile range, n≥8 in each group. Statistical analyses by Mann-Whitney U tests. Fluorescence data is presented by individual tetraspanin capture spots. Data from each tetraspanin capture spot (CD9, CD63 and CD81 – first three columns from the left) were calculated by subtracting negative control values from mouse IgG spots (shown in the furthest panel to the right). **A**) Monocyte-derived CD14^+^/CD81^+^ BAL EV were elevated in sepsis patients with ARDS compared to sepsis patients without ARDS (medians 1.23 ×10^8^/ml vs 6.26 ×10^7^/ml, p=0.015). In esophagectomy patients, those who subsequently developed ARDS had lower CD14^+^/CD81^+^ BAL EV counts compared to patients who did not develop ARDS (medians 3.96 ×10^7^/ml vs 9.94 ×10^7^/ml, p=0.03). **B**) Endothelial-derived CD31^+^/CD63^+^ and CD31^+^/CD81^+^ BAL EV were elevated in esophagectomy patients who subsequently developed ARDS compared to controls (p≤0.02). **C-F**) There were no differences in the numbers of CD66b^+^, CD41^+^, CD206^+^ and EpCam^+^ EV between ARDS and control groups in both cohorts.

In sepsis patients with ARDS, those who died within 30 days of ICU admission had a greater number of CD14^+^/CD81^+^ BAL EV than survivors (**Figure 3A**, medians 3.43 ×10^8^ /ml vs 9.54 ×10^7^ /ml, p=0.027), however there is overlap between groups. Across all sepsis patients with and without ARDS, CD66b^+^/CD63^+^ BAL EV correlated directly with BAL interleukin (IL)-8 concentrations, (**Figure 3B**, r=0.751, p<0.0001). Across all sepsis patients with and without ARDS, CD66b^+^/CD63^+^ BAL EV correlated inversely with alveolar macrophage efferocytosis index (**Figure 3C**, r=-0.612, p=0.0025).

**Figure 3:**
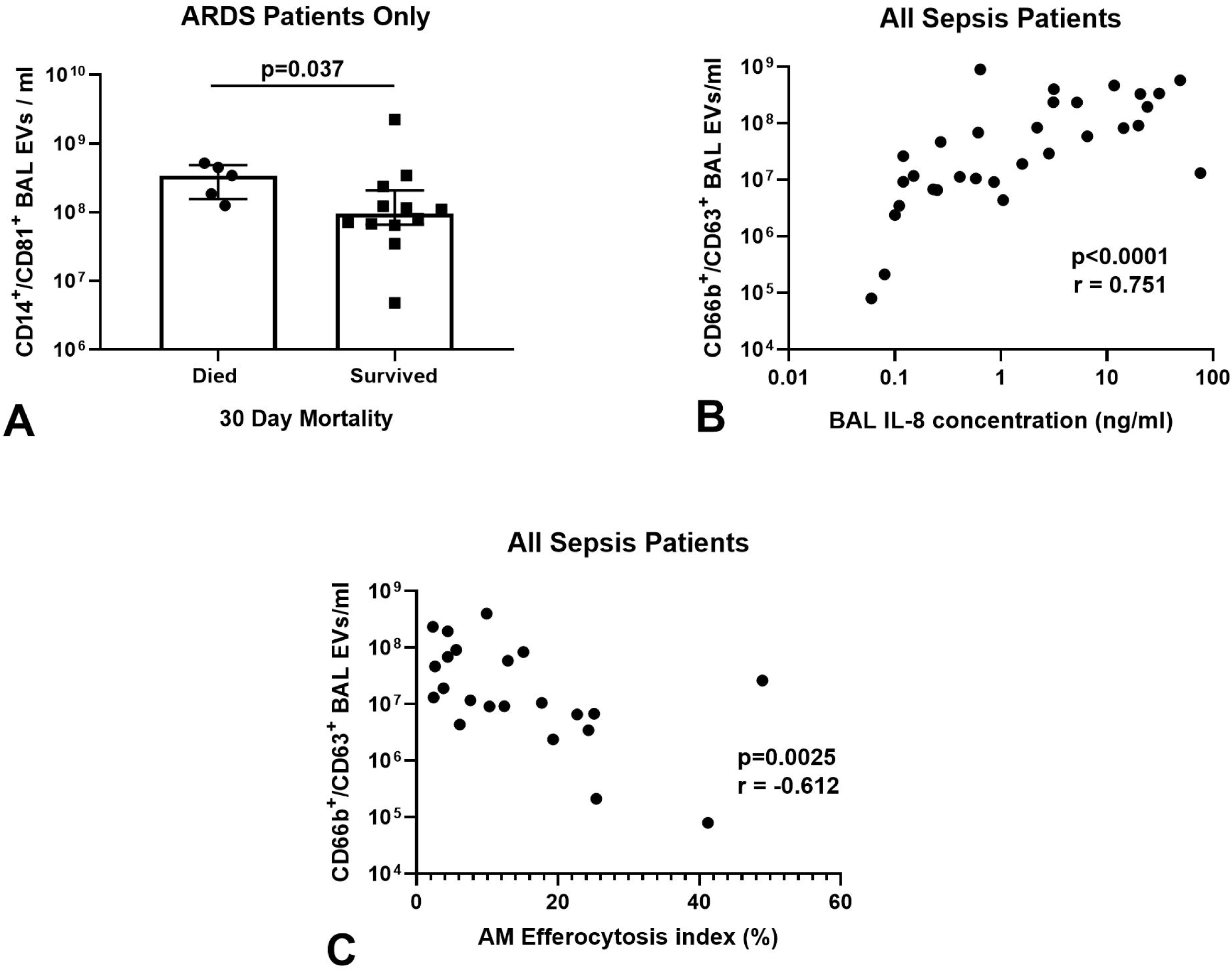
Correlations between BAL EV and clinical, biochemical and immune parameters. AM= Alveolar Macrophage. IL= Interleukin. **A**) In sepsis patients with ARDS, those who died within 30 days of ICU admission had a greater number of CD14^+^/ CD81^+^ BAL EV than survivors (medians 3.43 ×10^8^ /ml vs 9.54 ×10^7^ /ml, p=0.027, n=17). Statistical analysis by Mann-Whitney test, data shown as median and inter-quartile range. **B**) Across all sepsis patients, with and without ARDS, CD66b^+^/ CD63^+^ BAL EV correlate directly with BAL IL-8 concentrations (r=0.751, p<0.0001, n=31). Statistical analyses by Spearman’s correlation coefficient. **C**) Across all sepsis patients, with and without ARDS, CD66b^+^/CD63^+^ BAL EV correlate inversely with AM efferocytosis index (r=-0.612, p=0.0025, n=31). Statistical analyses by Spearman’s correlation coefficient.

## DISCUSSION

In this study, sepsis patients with ARDS had significantly higher numbers of CD14^+^/CD81^+^ BAL EV than sepsis patients without ARDS. CD14^+^/CD81^+^ BAL EV numbers were significantly higher in those patients with sepsis-related ARDS who died during the 30 days following ICU admission. CD206^+^ EV numbers were unchanged in sepsis-related ARDS patients (CD206 being an alveolar macrophage marker), indicating that the increased CD14^+^ EV observed in ARDS patients were derived from infiltrating monocytes as opposed to resident alveolar macrophages. CD14 has previously been used as a marker of monocyte-derived EV (18). The relationship between CD14^+^/CD81^+^ BAL EV and mortality in sepsis-related ARDS patients is in keeping with previous studies showing that the degree of monocyte influx in ARDS can correlate with the severity of respiratory failure (19). Previous studies have shown that monocyte-derived plasma EV containing gasdermin D and activated caspase-1 are more abundant in patients with sepsis-related ARDS compared to healthy controls; uptake of these EV can induce human pulmonary vascular endothelial cell death (20). This data supports our hypothesis that CD14^+^ EV may contribute to ARDS pathogenesis, and might be utilized as a biomarker of disease severity. Within the sepsis-related ARDS cohort, there appear to be two distinct subpopulations of BAL CD14^+^ EV numbers: ‘high’ and ‘low’ (Figure 2A). These may reflect different ARDS phenotypes (21), with the ‘high’ CD14^+^ EV subpopulation indicating a hyper-inflammatory phenotype, and the ‘low’ subpopulation indicating a hypo-inflammatory phenotype; however further analysis of BAL fluid and physiological data would be required to confirm this. Conversely, CD14^+^/CD81^+^ BAL EV counts were decreased in esophagectomy patients who later developed ARDS compared to controls. BAL samples from esophagectomy patients were taken immediately post-operatively prior to the development of ARDS, compared to being taken at the time of ARDS diagnosis in sepsis patients. This temporal difference in sampling and differences in the etiology of ARDS between the two cohorts may partially explain the divergent CD14^+^/CD81^+^ BAL EV counts. However, due to low n-numbers in the post-esophagectomy ARDS group, firm conclusions cannot be made and further appropriately powered studies are required to investigate the predictive role of CD14^+^/CD81^+^ BAL EV in esophagectomy patients.

We also observed associations between CD66b^+^/CD63^+^ neutrophil-derived BAL EV in sepsis patients and increased BAL IL-8 concentration and decreased alveolar macrophage efferocytosis. These data indicate that CD66b^+^/CD63^+^ BAL EV may be a marker of inflammation in sepsis-related ARDS. Neutrophil-derived EV have been shown to have an anti-inflammatory, tolerogenic effect following uptake by macrophages and T lymphocytes (22-25). Previous studies have also reported that neutrophil-derived EV can transfer microRNA-223 to alveolar epithelial cells, which reduces protein permeability and cytokine release via repression of poly[adenosine diphosphate–ribose] polymerase–1 (26). Therefore, the increased number of CD66b^+^ EV in patients with a more inflammatory alveolar microenvironment may be a compensatory protective response to suppress inflammation and reduce alveolar epithelial injury.

Our findings of increased CD31^+^/CD63^+^ and CD31^+^/CD81^+^ endothelial-derived EV in esophagectomy patients who subsequently developed ARDS may indicate an early signal of pulmonary endothelial injury resulting from one-lung ventilation detectable immediately post-operatively. This endothelial injury may initiate the development of ARDS in esophagectomy patients. Murine studies have shown that ventilator-induced lung injury can increase endothelial EV shedding (27); endothelial-derived EVs have been shown to induce acute lung injury by promoting neutrophil infiltration, endothelial dysfunction and pro-inflammatory cytokine release (28, 29). Appropriately powered future studies are required to further investigate the role of CD31^+^ EV as a predictive biomarker in esophagectomy patients. No difference in CD31^+^ and CD41^+^ EVs were observed between sepsis patients with and without ARDS. It is also important to note that the process of BAL dilutes the alveolar epithelial lining fluid by approximately 100-fold, which may account for BAL EV numbers being lower than EV numbers detected in other biofluids.

EV transfer of microRNA cargo to murine macrophages can alter metabolic profile and function (30); EV transfer of long non-coding RNA can promote glycolysis (31). An association between impaired alveolar macrophage efferocytosis and dependence on glycolysis has been observed in COPD patients (32). Further studies are required to determine if similar pathological processes occurs in ARDS, specifically whether CD14^+^ EV transfer of microRNA or protein cargo induces alveolar macrophage dysfunction (12), thereby promoting persistent inflammation. Understanding the role of EV in ARDS pathogenesis will allow identification of potential EV / microRNA biomarkers and novel therapeutic targets for ARDS.

A limitation of our study is that while we identified EV from the major cell types present in the alveolar space, not all cellular origins were included in our analysis (e.g. dendritic cells, lymphocytes). Although tetraspanin capture of EVs provides greater specificity compared to other isolation methods, it also limits phenotyping, and therefore only provides an indication of the relevance of CD14^+^ EV. Another limitation is that we did not capture EV based on CD14^+^, CD31^+^ and CD66b^+^ expression, which would allow probing of external markers for extensive phenotyping. Future studies are required to undertake this and probe for internal markers (e.g. syntenin) to determine EV biogenesis pathways. The finding that CD14 most commonly co-localises with tetraspanin CD81, and CD66b with tetraspanin CD63, may indicate different EV biogenesis pathways in monocytes and neutrophils (33). A further limitation of our study is that the observed differences in CD14^+^ BAL EVs were modest, and it is unclear whether these differences are biologically relevant. Similarly, correlations observed with CD14^+^, CD31^+^ and CD66b^+^ BAL EVs cannot be used to draw conclusions on their functional roles. Further work is required to determine the biological function of CD14^+^ and CD31^+^ BAL EV subpopulations in ARDS patients. Another limitation of our study is that the number of VINDALOO trial patients who received placebo and developed ARDS following esophagectomy was low (n=4), thus preventing us from drawing firm conclusions regarding the role of EVs as biomarkers in this population.

In conclusion, we report that CD14^+^/CD81^+^ BAL EV are enriched in patients with sepsis–related ARDS compared to controls, and that an elevated CD14^+^/CD81^+^ BAL EV count is associated with increased mortality in ARDS patients, although the sample size is modest. Thus, CD14^+^ BAL EV are a potential biomarker for disease severity and mortality in ARDS. Our findings provide the impetus to further elucidate the role these EV play in ARDS pathogenesis.

## Data Availability

Anonymised data are available upon reasonable request to the corresponding author.

## Ethical Approval

Ethical approval was obtained to recruit ventilated sepsis patients with and without ARDS (REC 16/WA/0169). Ethical approval was also obtained to recruit patients undergoing transthoracic esophagectomy for esophageal carcinoma (REC 12/WM/0092) as previously described in the VINDALOO trial. For patients who lacked capacity, permission to enroll was sought from a personal legal representative in accordance with the UK Mental Capacity Act (2005). For patients with capacity, written informed consent was obtained from the patient.

## Grants

This work was funded by Medical Research Council grants MR/N021185/1 (RYM) and MR/L002736/1 (DT/AS), and by National Heart, Lung and Blood Institute grants HL134828 (MAM) and HL140026 (MAM). The ExoView platform was funded by an EPSRC Capital award awarded to Dr. Sophie Cox, University of Birmingham.

## Disclosures

No conflicts of interest, financial or otherwise, are declared by the authors.

## Author Contributions

RYM, AS, DP, PH, MAM, and DRT contributed to study conception and design. RYM, JP, HL and STL contributed to data acquisition. JP and PH provided assistance and training for use of the Exoview platform. All authors contributed to the data analysis and interpretation. RYM, JP, and DRT drafted the manuscript. All authors critically revised the manuscript for intellectual content and approved the final version before submission.

## Notes

### Competing Interest Statement

The authors have declared no competing interest.

### Author Declarations

Ethical approval was given by the Wales Research Ethics Committee 1 (reference number 16/WA/0169) to recruit ventilated sepsis patients with and without ARDS. Ethical approval was also given by the South Birmingham (England) Research Ethics Committee (reference number 12/WM/0092) to recruit patients undergoing transthoracic esophagectomy for esophageal carcinoma as previously described in the VINDALOO trial. For patients who lacked capacity, permission to enroll was sought from a personal legal representative in accordance with the UK Mental Capacity Act (2005). For patients with capacity, written informed consent was obtained from the patient.

### Summary of Updates

We have now included additional analyses performed on a subset of esophagectomy patients from the VINDALOO trial who subsequently developed ARDS. We have also altered our statistical analyses to undertake internal comparisons within the sepsis patient cohort and the esophagectomy patient cohort, and removed comparisons between sepsis and esophagectomy patient cohorts.

## REFERENCES

1. Matthay MA, Zemans RL, Zimmerman GA, Arabi YM, Beitler JR, Mercat A, Herridge M, Randolph AG, and Calfee CS. Acute respiratory distress syndrome. Nature reviews Disease primers 5: 18, 2019.

2. Bellani G, Laffey JG, Pham T, Fan E, Brochard L, Esteban A, Gattinoni L, van Haren F, Larsson A, McAuley DF, Ranieri M, Rubenfeld G, Thompson BT, Wrigge H, Slutsky AS, and Pesenti A. Epidemiology, Patterns of Care, and Mortality for Patients With Acute Respiratory Distress Syndrome in Intensive Care Units in 50 Countries. Jama 315: 788–800, 2016.

3. Yang X, Yu Y, Xu J, Shu H, Xia J, Liu H, Wu Y, Zhang L, Yu Z, Fang M, Yu T, Wang Y, Pan S, Zou X, Yuan S, and Shang Y. Clinical course and outcomes of critically ill patients with SARS-CoV-2 pneumonia in Wuhan, China: a single-centered, retrospective, observational study. The Lancet Respiratory medicine 8: 475–481, 2020.

4. Mahida RY, Matsumoto S, and Matthay MA. Extracellular Vesicles: A New Frontier for Research in Acute Respiratory Distress Syndrome. American journal of respiratory cell and molecular biology 63: 15–24, 2020.

5. Shah R, Patel T, and Freedman JE. Circulating Extracellular Vesicles in Human Disease. The New England journal of medicine 379: 958–966, 2018.

6. Gupta R, Radicioni G, Abdelwahab S, Dang H, Carpenter J, Chua M, Mieczkowski PA, Sheridan JT, Randell SH, and Kesimer M. Intercellular Communication between Airway Epithelial Cells Is Mediated by Exosome-Like Vesicles. American journal of respiratory cell and molecular biology 60: 209–220, 2019.

7. Khan NZ, Cao T, He J, Ritzel RM, Li Y, Henry RJ, Colson C, Stoica BA, Faden AI, and Wu J. Spinal cord injury alters microRNA and CD81+ exosome levels in plasma extracellular nanoparticles with neuroinflammatory potential. Brain, behavior, and immunity 92: 165–183, 2021.

8. Kugeratski FG, Hodge K, Lilla S, McAndrews KM, Zhou X, Hwang RF, Zanivan S, and Kalluri R. Quantitative proteomics identifies the core proteome of exosomes with syntenin-1 as the highest abundant protein and a putative universal biomarker. Nature Cell Biology 23: 631–641, 2021.

9. Liu A, Park JH, Zhang X, Sugita S, Naito Y, Lee JH, Kato H, Hao Q, Matthay MA, and Lee JW. Therapeutic Effects of Hyaluronic Acid in Bacterial Pneumonia in the Ex Vivo Perfused Human Lungs. American journal of respiratory and critical care medicine 2019.

10. Shikano S, Gon Y, Maruoka S, Shimizu T, Kozu Y, Iida Y, Hikichi M, Takahashi M, Okamoto S, Tsuya K, Fukuda A, Mizumura K, and Hashimoto S. Increased extracellular vesicle miRNA-466 family in the bronchoalveolar lavage fluid as a precipitating factor of ARDS. BMC pulmonary medicine 19: 110, 2019.

11. Bazzan E, Radu CM, Tinè M, Neri T, Biondini D, Semenzato U, Casara A, Balestro E, Simioni P, Celi A, Cosio MG, and Saetta M. Microvesicles in bronchoalveolar lavage as a potential biomarker of COPD. American journal of physiology Lung cellular and molecular physiology 320: L241–l245, 2021.

12. Mahida RY, Scott A, Parekh D, Lugg ST, Hardy RS, Lavery GG, Matthay MA, Naidu B, Perkins GD, and Thickett DR. Acute Respiratory Distress Syndrome is associated with impaired alveolar macrophage efferocytosis. The European respiratory journal 2021.

13. Mahida RY, Scott A, Parekh D, Lugg ST, Belchamber KB, Hardy RS, Matthay MA, Naidu B, and Thickett DR. Assessment of alveolar macrophage dysfunction using an in vitro model of Acute Respiratory Distress Syndrome. Front Med (Lausanne) In Press., 2021.

14. Parekh D, Dancer RC, Lax S, Cooper MS, Martineau AR, Fraser WD, Tucker O, Alderson D, Perkins GD, Gao-Smith F, and Thickett DR. Vitamin D to prevent acute lung injury following oesophagectomy (VINDALOO): study protocol for a randomised placebo controlled trial. Trials 14: 100, 2013.

15. Parekh D, Dancer RCA, Scott A, D’Souza VK, Howells PA, Mahida RY, Tang JCY, Cooper MS, Fraser WD, Tan L, Gao F, Martineau AR, Tucker O, Perkins GD, and Thickett DR. Vitamin D to Prevent Lung Injury Following Esophagectomy-A Randomized, Placebo-Controlled Trial. Critical care medicine 46: e1128–e1135, 2018.

16. Haslam PL, and Baughman RP. Report of ERS Task Force: guidelines for measurement of acellular components and standardization of BAL. The European respiratory journal 14: 245–248, 1999.

17. Price J, Gardiner C, and Harrison P. Platelet-enhanced plasma: Characterization of a novel candidate resuscitation fluid’s extracellular vesicle content, clotting parameters, and thrombin generation capacity. Transfusion 61: 2179–2194, 2021.

18. Wang GH, Lu J, Ma KL, Zhang Y, Hu ZB, Chen PP, Lu CC, Zhang XL, and Liu BC. The Release of Monocyte-Derived Tissue Factor-Positive Microparticles Contributes to a Hypercoagulable State in Idiopathic Membranous Nephropathy. Journal of atherosclerosis and thrombosis 26: 538–546, 2019.

19. Rosseau S, Hammerl P, Maus U, Walmrath HD, Schutte H, Grimminger F, Seeger W, and Lohmeyer J. Phenotypic characterization of alveolar monocyte recruitment in acute respiratory distress syndrome. American journal of physiology Lung cellular and molecular physiology 279: L25–35, 2000.

20. Mitra S, Exline M, Habyarimana F, Gavrilin MA, Baker PJ, Masters SL, Wewers MD, and Sarkar A. Microparticulate Caspase 1 Regulates Gasdermin D and Pulmonary Vascular Endothelial Cell Injury. American journal of respiratory cell and molecular biology 59: 56–64, 2018.

21. Calfee CS, Delucchi K, Parsons PE, Thompson BT, Ware LB, and Matthay MA. Subphenotypes in acute respiratory distress syndrome: latent class analysis of data from two randomised controlled trials. The Lancet Respiratory medicine 2: 611–620, 2014.

22. Shen G, Krienke S, Schiller P, Nießen A, Neu S, Eckstein V, Schiller M, Lorenz HM, and Tykocinski LO. Microvesicles released by apoptotic human neutrophils suppress proliferation and IL-2/IL-2 receptor expression of resting T helper cells. European journal of immunology 47: 900–910, 2017.

23. Eken C, Martin PJ, Sadallah S, Treves S, Schaller M, and Schifferli JA. Ectosomes released by polymorphonuclear neutrophils induce a MerTK-dependent anti-inflammatory pathway in macrophages. The Journal of biological chemistry 285: 39914–39921, 2010.

24. Gasser O, and Schifferli JA. Activated polymorphonuclear neutrophils disseminate anti-inflammatory microparticles by ectocytosis. Blood 104: 2543–2548, 2004.

25. Dengler V, Downey GP, Tuder RM, Eltzschig HK, and Schmidt EP. Neutrophil intercellular communication in acute lung injury. Emerging roles of microparticles and gap junctions. American journal of respiratory cell and molecular biology 49: 1–5, 2013.

26. Neudecker V, Brodsky KS, Clambey ET, Schmidt EP, Packard TA, Davenport B, Standiford TJ, Weng T, Fletcher AA, Barthel L, Masterson JC, Furuta GT, Cai C, Blackburn MR, Ginde AA, Graner MW, Janssen WJ, Zemans RL, Evans CM, Burnham EL, Homann D, Moss M, Kreth S, Zacharowski K, Henson PM, and Eltzschig HK. Neutrophil transfer of miR-223 to lung epithelial cells dampens acute lung injury in mice. Science translational medicine 9: 2017.

27. Letsiou E, Sammani S, Zhang W, Zhou T, Quijada H, Moreno-Vinasco L, Dudek SM, and Garcia JG. Pathologic mechanical stress and endotoxin exposure increases lung endothelial microparticle shedding. American journal of respiratory cell and molecular biology 52: 193–204, 2015.

28. Buesing KL, Densmore JC, Kaul S, Pritchard KA, Jr., Jarzembowski JA, Gourlay DM, and Oldham KT. Endothelial microparticles induce inflammation in acute lung injury. The Journal of surgical research 166: 32–39, 2011.

29. Densmore JC, Signorino PR, Ou J, Hatoum OA, Rowe JJ, Shi Y, Kaul S, Jones DW, Sabina RE, Pritchard KA, Jr., Guice KS, and Oldham KT. Endothelium-derived microparticles induce endothelial dysfunction and acute lung injury. Shock (Augusta, Ga) 26: 464–471, 2006.

30. Park JE, Dutta B, Tse SW, Gupta N, Tan CF, Low JK, Yeoh KW, Kon OL, Tam JP, and Sze SK. Hypoxia-induced tumor exosomes promote M2-like macrophage polarization of infiltrating myeloid cells and microRNA-mediated metabolic shift. Oncogene 38: 5158–5173, 2019.

31. Chen F, Chen J, Yang L, Liu J, Zhang X, Zhang Y, Tu Q, Yin D, Lin D, Wong PP, Huang D, Xing Y, Zhao J, Li M, Liu Q, Su F, Su S, and Song E. Extracellular vesicle-packaged HIF-1α-stabilizing lncRNA from tumour-associated macrophages regulates aerobic glycolysis of breast cancer cells. Nature cell biology 21: 498–510, 2019.

32. Ryan E, Coelho P, Cole J, Bewley M, Budd R, Callahan J, McCafferty J, Singh D, Dockrell D, Walmsley S, and Whyte M. T1 Defective metabolism drives macrophage dysfunction in COPD. Thorax 76: A1–A1, 2021.

33. Mathieu M, Névo N, Jouve M, Valenzuela JI, Maurin M, Verweij FJ, Palmulli R, Lankar D, Dingli F, Loew D, Rubinstein E, Boncompain G, Perez F, and Théry C. Specificities of exosome versus small ectosome secretion revealed by live intracellular tracking of CD63 and CD9. Nature communications 12: 4389, 2021.

